# Subsequent hip fractures in patients identified by a Fracture Liaison Service (FLS) in England and Wales – linkage of the national FLS and Hip Fracture databases

**DOI:** 10.1101/2022.01.10.22268911

**Authors:** MK Javaid, Z Mohsin, A Johansen, CL Gregson, R Pinedo-Villanueva

## Abstract

Fracture Liaison Services (FLSs) are recommended healthcare models to deliver secondary fracture prevention and reduce the risk of subsequent fractures. Several studies have demonstrated the cost and clinical effectiveness of FLSs, but there is little real-world data on the impact of FLSs on subsequent hip fracture rates.

A cohort of 50,214 patients from the national FLS database with an index fracture of the hip, spine, or other in 2017 was linked to the National Hip Fracture Database from 2017 to 2020 to identify those patients who went on to have a subsequent hip fracture.

One in twenty (5.1%) of the 9,888 people in whom the index fracture was at the hip went on to suffer a second hip fracture within 3-4 years, despite receiving the support of an FLS. The risk of hip fracture was similar (4.7%) if the index fracture was at the spine, but lower at other sites (2.8%, p<0.001) and the interval shortest after an index hip fracture (1.1 years (0.4,2.0) p<0.001). The proportion of patients with a subsequent hip fracture was not lower by types of anti-osteoporotic medication.

This work highlights the need for alternative anti-osteoporotic management strategies to rapidly decrease the risk of subsequent hip fractures for people seen by an FLS setting with levels of risk that are even higher for patients in areas which are still not served by an FLS.

## Introduction

Approximately 500,000 adults sustain a fragility fracture every year in the UK[1]. This is often the first sign of osteoporosis and a critical opportunity for the patient, healthcare system and society to intervene to improve bone health, reduce the risk of future fractures, and avoid further injury. In 2009, the NHS prioritised Fracture Liaison Services (FLSs) to provide osteoporosis and falls checks for patients after a fragility fracture. There are established methods to identify patients and assess fracture risk[2], and decide which patients require therapy to reduce fracture risk[3, 4]. Several studies and reviews have demonstrated the value of FLSs to reduce fracture risk compared to usual care[5-7]. We have previously demonstrated that not all FLSs are automatically effective, for example no reduction in re-fracture rates were seen in patients post hip fracture, using an interrupted time series analysis in 11 NHS hospital trusts[8]. A qualitative study to understand the reasons behind this finding identified the importance for an FLS to monitor patients after recommending treatment to ensure initiation and adherence to treatment, a component of FLS pathway that was often delegated to primary care[9]. Adherence to oral bisphosphonates adherence after a hip fracture using real-world data, confirms poor treatment adherence[10]. These data have informed the development of FLS clinical standards by the Royal Osteoporosis Society[11] that include longer term follow-up to optimise adherence and the need to embed quality improvement within FLSs so they can deliver the expected reduction in fracture risk.

In 2016, to support effective delivery of FLSs, the Healthcare Quality Improvement Partnership, funded by NHS England and Welsh Governments, commissioned the world’s first mandatory audit of FLSs: the Fracture Liaison Service Database (FLSDB) audit of England and Wales[12]. The FLSDB multidisciplinary advisory group, including patients and patient representatives, developed 11 outcome and process performance indicators covering identification, assessment, treatment recommendation and adherence[13] that have since informed the global FLS key performance indicator set[14]. Since its inception, the FLSDB has demonstrated increasing participation and delivery of key performance indicators through a series of annual reports[13, 15-17]. However, the impact of the audit on patients’ outcomes is not known. In this study we describe the incident rates of subsequent hip fracture in patients who have been managed by an FLS.

## Materials and Methods

### Data sources, data linkage and selection of study population

Patient records from two national clinical audits, and the Fracture Liaison Service Database for England and Wales (FLSDB)[16] in 2017 (1^st^ January to 31^st^ December) and the National Hip Fracture Database for England and Wales (NHFD)[18] from 1^st^ Jan 2016 to 31^st^ December 2020, were provided using patient-level pseudo-anonymised identifiers. The content for the FLSDB and NHFD are available online (https://www.fffap.org.uk/fls/flsweb.nsf/ and nhfd.co.uk). The data for this analysis was collected under Regulation 5 of the Health Service (Control of Patient Information) Regulations 2002 to process identifiable information without consent (reference 15/CAG/0158). Approval for using these data for this study was granted by the chair of the Scientific and Publications Committee, Falls and Fragility Fracture Audit Programme, the Falls and Fragility Fracture Audit Programme Deputy Programme Manager as data provider and Health Quality Improvement Partnership as data controllers (reference HQIP386).

The FLSDB records fracture site in three groups: hip, spine and other. Where there was more than one fracture site recorded on the same day, then the index fracture site was recorded as hip, then spine then other. Records without a skeletal site for their index fracture were excluded (S1 Fig.). If more than one fracture was recorded within the FLSDB for the same patient in 2017, we used the earliest record as their index fracture date and site, resulting in a total of 50,214 index FLSDB fracture records.

Within the NHFD dataset from 2016 to 2020, we excluded fracture locations other than hip (femoral shaft, distal femoral and peri-prosthetic fractures) and then linked the remaining records of hip fractures with the 50,214 index cases from the FLSDB in 2017. S1 Ta describes the number of NHFD hip fractures linked to an index FLDSB fracture record, arranged by the time interval between the two fracture records. An index hip fracture recorded in the FLSDB might also have been entered into NHFD, so to avoid double counting, we excluded NHFD hip fracture records with a presentation date that preceded or was less than 30 days after an FLSDB index hip fracture in the FLSDB in 2017. Since the focus of this study was on the potential for fracture prevention and these very early events were considered unlikely to be preventable by any FLS intervention. We excluded NHFD hip fracture records within 30 days of a FLSDB record. Where more than one hip fracture occurred after an index FLSDB fracture, the earliest hip fracture in the NHFD was analysed.

### Predictors of incident hip fracture

We examined FLSDB records of age, sex and fracture site, as well as anti-osteoporosis medication (AOM) prescriptions at the time of index fracture (i.e. fracture on treatment). Sixty-six patients (0.1%) had more than one type of AOM recorded at the time of index fracture. We used the following hierarchy to assign the AOM recorded at the time of index fracture: teriparatide, denosumab, zoledronate, other oral AOM (raloxifene, oestrogen hormone therapy, strontium ranelate, alfacalcidol, calcitriol), then oral bisphosphonates. Similarly, 850 (1.7%) of FLSDB records had a treatment recommendation more than one type of AOM. Given prescribers would in general use the least expensive treatment recommended[3], the likely AOM recommended post FLS assessment were assigned using the following hierarchy: oral bisphosphonates, other oral AOMs (raloxifene, oestrogen hormone therapy, strontium ranelate, alfacalcidol, calcitriol), zoledronate, denosumab, and then teriparatide.

### Statistical Analysis

Descriptive parametric and non-parametric tests were used to compare baseline characteristics between relevant groups. As, the FLSDB dataset does not include subsequent mortality, we were unable to adjust for the competing risk of death or survival bias. Stata/MP 15.1 software (StataCorp, TX, USA) was used with p<0.05 to indicate statistical significance.

## Results

We identified 50,214 patients from the FLSBD with at least one index fracture record in 2017. Their baseline characteristics are shown in Table 1 by site of index fracture. Hip fractures made up 19.7% of index fracture records. Compared with the cohort of hip fractures in the NHFD in 2017 (n=64,625), the patients a hip fractures in the FLSDB were more likely to be younger (80.6 vs 82.8 years, p<0.001) with a similar proportion female (71.2% vs 70.7%, p=0.09).

**Table 1:** Baseline characteristics of the records from the FLSDB with an index fragility fracture in 2017.

| Characteristic |  | Index FLSDb recorded fracture site |  |  |
| --- | --- | --- | --- | --- |
|  |  | Hip | Spine | Other |
| n |  | 9,886 | 2,985 | 37,341 |
| Female(%) |  | 7,036 (71.2%) | 2,088 (70.0%) | 30,128 (80.7%) |
| Age years | Mean (SD) | 80.6 (10.3) | 74.9 (10.5) | 69.8 (11.7) |
| Height(m) | Mean (SD) | 1.64 (0.10) | 1.62 (0.10) | 1.63 (0.09) |
|  | Missing | 5,179 (52.4%) | 1,242 (41.6%) | 13,909 (37.3%) |
| Weight(kg) | Median (IQR) | 64.0 (55, 74.8) | 67.1 (58, 78.8) | 70 (60.3, 82) |
|  | Missing | 5,180 (52.4%) | 1,185 (39.7%) | 13,111 (35.1%) |
| BMI(kg/m<sup>2</sup>) | Median (IQR) | 23.8 (21.0, 26.8) | 25.9 (22.8,29.4) | 26.3 (23.4, 30.3) |
|  | Missing | 5,301 (53.6%) | 1,256 (42.1%) | 14,313 (38.3%) |
| Care home resident pre-fracture | Yes | 1,420 (14.4%) | 85 (2.9%) | 1,328 (3.6%) |
|  | Missing | 321 (3.3%) | 185 (6.2%) | 1,145 (3.1%) |
| Previous fracture (%) | Yes | 2,111(21.4%) | 1,096 (36.7%) | 9,793(26.2%) |
|  | Missing | 2,495 (25.2%) | 564 (18.9%) | 7,808 (20.9%) |
| Parental hip fracture (%) | Yes | 316 (3.2%) | 213 (7.1%) | 2,923(7.8%) |
|  | Missing | 2,871(29.0%) | 631 (21.2%) | 7,876(21.1%) |
| Current smoker(%) | Yes | 659 (6.7%) | 244(8.2%) | 2,991 (8.0%) |
|  | Missing | 2,403(24.3%) | 602 (20.2%) | 7,473 (20.0%) |
| Previous AOM therapy | Unknown | 2,601 (26.3%) | 652 (21.8%) | 8,752 (23.4%) |
|  | No | 6,383 (64.6%) | 1,905(63.8%) | 26,016 (69.7%) |
|  | Oral BP<sup>1</sup> | 729 (7.4%) | 365 (12.2%) | 2,052 (5.5%) |
|  | Other AOM<sup>2</sup> | 33 (0.3%) | 11 (0.4%) | 217 (0.6%) |
|  | Zoledronate | 46 (0.5%) | 26 (0.9%) | 140 (0.4%) |
|  | Denosumab | 88 (0.9%) | 24 (0.8%) | 156 (0.4%) |
|  | Teriparatide | 5 (0.05%) | 2(0.07%) | 10(0.03%) |
| Bone mineral testing using DXA | Not ordered | 6,261 (63.3%) | 1,098 (36.8%) | 11,459 (30.7%) |
|  | Done in last 2 years | 401 (4.1%) | 301 (10.1%) | 2,383 (6.4%) |
|  | Ordered/recommended | 2,310 (23.4%) | 1,328 (44.5%) | 20,766 (55.6%) |
|  | Missing | 914 (9.2%) | 258 (8.6%) | 2735 (7.3%) |
|  | Result (n) | 625(34.5%) | 1,575(53.6%) | 20,425(55.4%) |
|  | Lowest T score<sup>3</sup>: |  |  |  |
| | T score $\leq -2.5$ | 920 (41.2%) | 779 (49.0%) | 5,201 (25.3%) |
|  | -2.5 to -1 | 905 (40.6%) | 586 (36.8%) | 9,817 (47.8%) |
| | $>-1$ | 406 (18.2%) | 226 (14.2%) | 5,543 (27.0%) |

Legend: <sup>1</sup> oral bisphosphonates include: alendronate, risedronate, ibandronate, etidronate; <sup>2</sup> other AOMs include raloxifene, strontium ranelate, alfacalcidol and oestrogen hormone therapy. <sup>3</sup>In those with reported DXA scores

Approximately a quarter (25.9%) of patients with an index fracture in 2017 were recorded to have a previous history of fragility fracture. While these patients were more likely to be on an AOM at the time of the index fracture than those without a history of previous fracture (17.6% vs 5.0%, p<0.001), it still represents a relatively small proportion on treatment.

The outcomes of the FLS assessment are shown in Table 2. Bone density assessment had been ordered or recommended in 49.6% of all patients, including 23.4% patients with an index hip fracture. Across all fracture sites, 26,430 (52.6%) patients were recommended AOM or referred to another clinician for consideration of bone health management. Oral bisphosphonates were the commonest AOM recommended (n=9,847) followed by subcutaneous denosumab (n=1,708) and intravenous zoledronate (n=1,637).

**Table 2:**
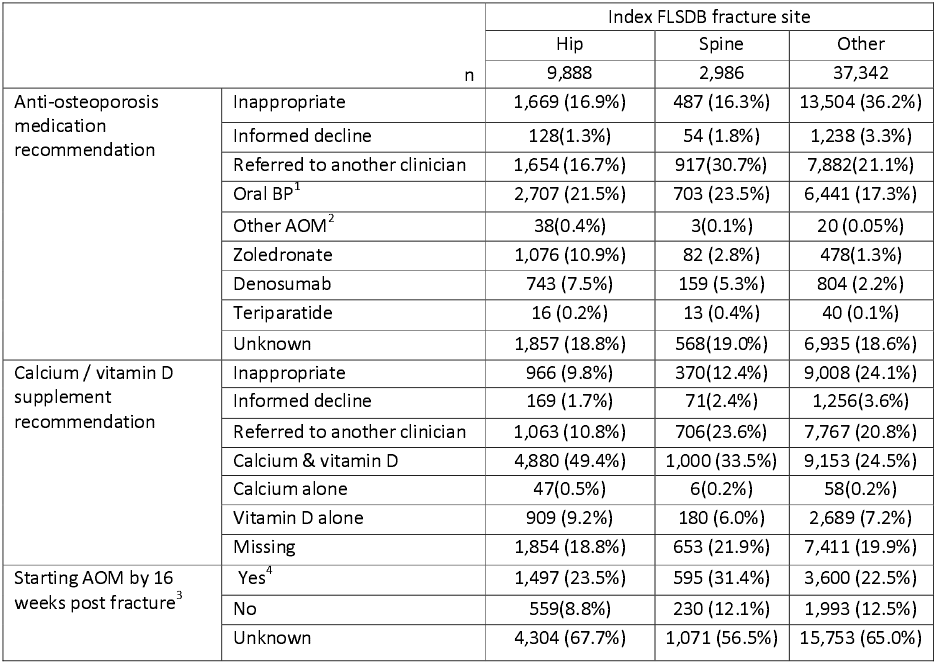
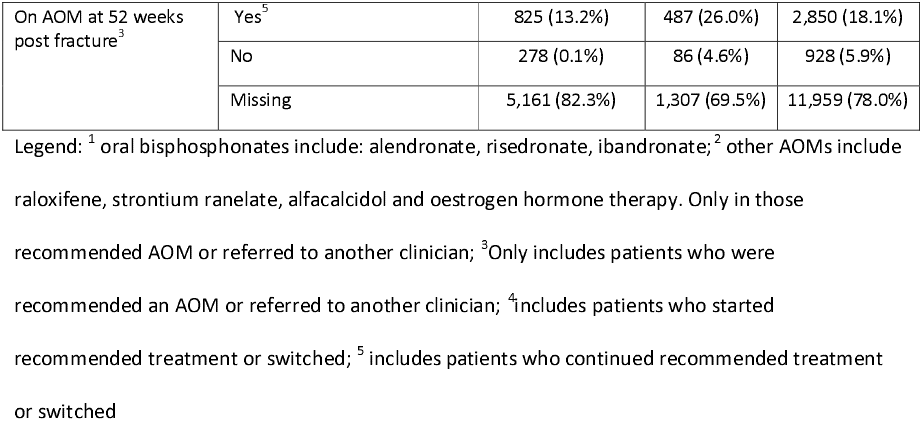
FLS outcomes by Fracture site.

In total 1,695 patients (3.4%) went on to have a subsequent hip fracture recorded in the NHFD by the end of 2020 (Table 3). The median time to subsequent hip fracture as 1.4 years (IQR 0.6, 2.3), with shorter intervals (0<0.001) for hip compared to spine or other index fracture sites. Patients who had a subsequent hip fracture were also older (81.2 vs 71.9 years, p<0.001) and more likely to be female (3.5% vs 3.1%, p=0.04). Patients with an index hip fracture were the most likely group to have a subsequent further hip fracture recorded in the NHFD (hip (5.1%), spine (4.7%) vs other sites (2.8%, p<0.001). In patients with an index hip fracture, both age and the proportion that were female varied significantly (p<0.001) by treatment recommendation (Table 4) as did the proportion of patients who subsequently fracture their hip fracture by type of FLS treatment recommendation for all patients (p=0.02) (Table 4) and by sex in Fig 1.

**Table 3:** Record of hip fracture in the NHFD at least 30 days after the index fracture in FLSDB.

| Index FLSDb fracture site in 2017 |  |  |  |
| --- | --- | --- | --- |
|  | Hip | Spine | Other |
| n | 9,888 | 2,986 | 37,342 |
| Years to subsequent hip fracture (median (IQR))<sup>1</sup> |  |  |  |
|  | 1.1 (0.4, 2.0) | 1.4 (0.6, 2.3) | 1.5 (0.8, 2.5) |
| Year a subsequent hip was fracture recorded in NHFD |  |  |  |
| 2017 | 148(28.9%) | 22(15.8%) | 167(16.0%) |
| 2018 | 155(30.3%) | 51(36.7%) | 362(34.7%) |
| 2019 | 133(26.0%) | 35(25.2%) | 267(25.6%) |
| 2020 | 76(14.8%) | 31(22.3%) | 248(23.8%) |
| **Total** | **512 (5.2%)** | **139 (4.7%)** | **1,044 (2.8%)** |
|  |  |  | p<0.001 |

Legend: column percentages are shown. Significance shown using Chi<sup>2</sup> tests. <sup>1</sup>Note this interval includes the exclusion of subsequent hip fractures within 30 days of index FLSDB patient record.

**Table 4:** Subsequent hip fracture by FLS treatment recommendation following index FLSDB hip fracture.

|  | Index FLSDb fracture site = hip |  |  |  |  |  |  |  |  |
| --- | --- | --- | --- | --- | --- | --- | --- | --- | --- |
|  | Unknown/missing | Inappropriate | Informed decline | Referred<sup>1</sup> | Oral BP<sup>2</sup> | Other AOM<sup>3</sup> | Zol<sup>4</sup> | Dmb<sup>5</sup> | PTH<sup>6</sup> |
| n | 1,857 | 1,669 | 128 | 1,654 | 2,707 | 38 | 1,076 | 744 | 16 |
| Female (%) | 68.5% | 65.4% | 66.4% | 69.5% | 75.6% | 52.6% | 71.7% | 76.8% | 100% |
| Mean age (SD) | 80.0 (10.5) | 80.5 (11.8) | 75.6 (11.8) | 77.6 (11.1) | 80.5 (9.3) | 85.2 (10.0) | 84.7 (8.0) | 83.4 (7.8) | 76.7 (8.0) |
|  | Subsequent hip fracture recorded in NHFD |  |  |  |  |  |  |  |  |
| 2017 | 30 | 24 | 2 | 20 | 37 |  | 20 | 15 | 0 |
| 2018 | 28 | 23 | 2 | 24 | 43 |  | 20 | 15 | 0 |
| 2019 | 25 | 11 | 0 | 26 | 37 |  | 14 | 19 | 1 |
| 2020 | 13 | 10 | 0 | 15 | 22 |  | 9 | 8 | 0 |
| **Total** | 95 (5.1%) | 68 (4.1%) | 4 (3.1%) | 85 (5.1%) | 139 (5.1%) | 0 | 63 (5.9%) | 55 (7.6%) | 1 (6.3%) |

Legend: <sup>1</sup>Referred = referred to other clinician/ GP; <sup>2</sup>Oral BP: alendronate, risedronate, ibandronate; <sup>3</sup> Other AOMs include raloxifene, strontium ranelate, alfacalcidol and oestrogen hormone therapy; <sup>4</sup> Zol= zoledronate; <sup>5</sup>Dmb= denosumab; <sup>6</sup>PTH= PTH analogues including teriparatide. Chi<sup>2</sup> test performed excluding informed decline, other AOM and PTH due to small numbers

**Fig 1.**
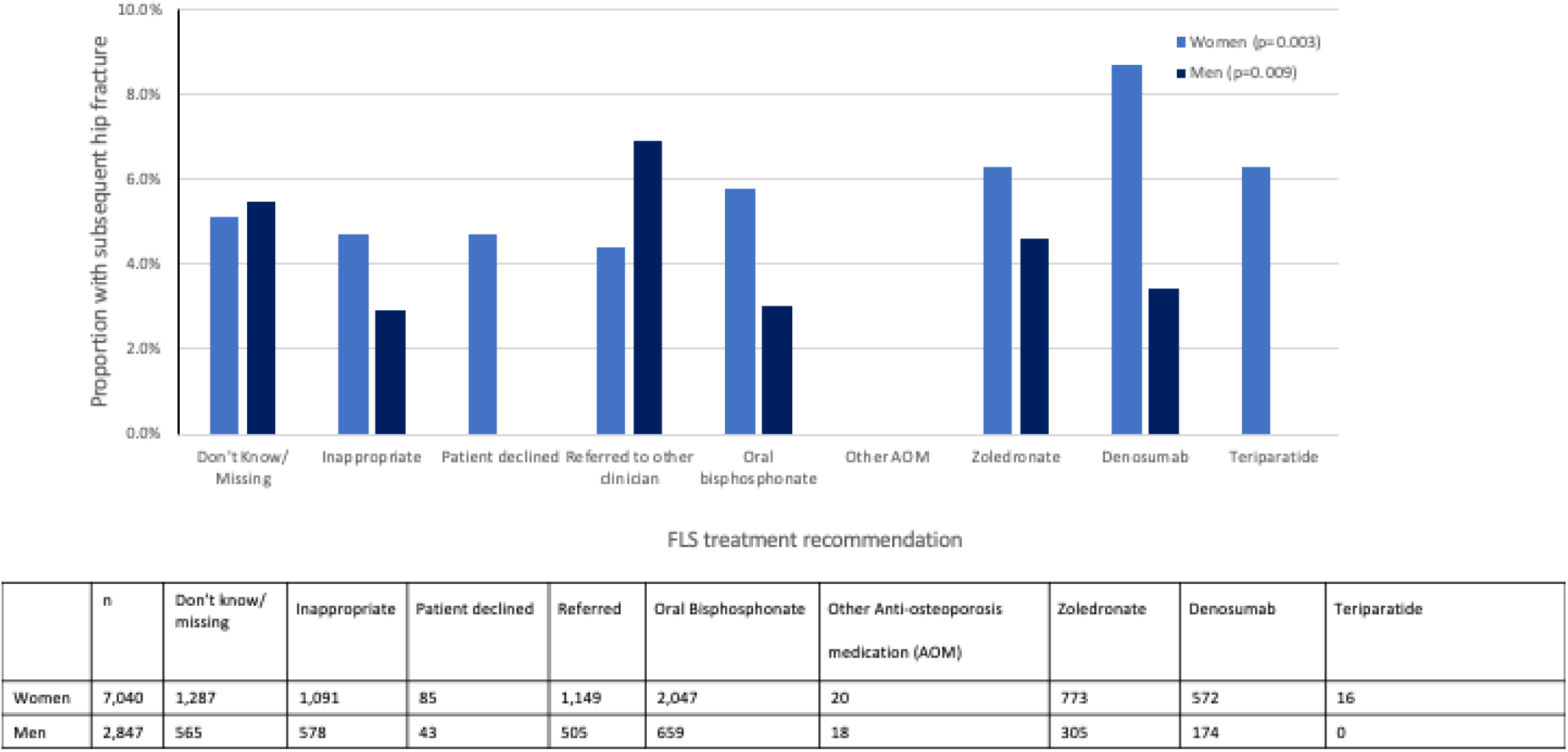
Proportion with a subsequent hip fracture by anti-osteoporosis medication recommended by the FLS, in patients with an index hip fracture by sex. Legend: The proportion with a subsequent hip fracture as recorded by the NHFD are shown by treatment recommended by the FLS by sex. The panel below shows the denominator for each treatment recommended by sex

## Discussion

We have demonstrated that despite receiving the support of FLS management one in twenty people (5.1%) who sustain one hip fracture will go on to suffer the second one within 3 to 4 years. Whilst we were unable to adjust for confounding by indication, survival bias or exclude those who with bilateral hip fractures at baseline, these findings suggest that current pathways within FLSs may be insufficient to substantially reduce imminent hip fracture risk in patients with a recent fracture especially at the hip and spine. Preventing a second hip fracture is important as mortality after a second hip fracture is significantly higher than after the first hip fracture[19] with reduction in physical, functional, and mental wellbeing outcomes, as well as a potentially avoidable burden for patients, their families, healthcare systems, and society[20]. While reviews of the literature have indicated that pharmacological therapy reduces the risk of a second hip fracture[21], these analyses suggest the residual risk of a hip fracture is substantial, especially in the 2 years following an index fragility fracture. This inference is supported by simulation models that account for imminent fracture risk, time to treatment effect, and scale of treatment effect [22].

A Canadian study of 532 FLS patients followed up over 2 years had a subsequent fracture rate of 2.6%, with a subsequent hip fracture rate of 0.1% [23]. A larger study from Hong Kong of 41,433 primary hip fractures has reported a cumulative incidence of 1.24% at 1 year and 4.42% at 5 years. Data comparing primary and secondary healthcare records from the UK, Catalonia and Denmark reported one-year incidence rates for second hip fracture per 1000 person years of 14.6, 11.6 and 37.9 respectively[24]. These reported rates are lower than that observed in this study and may be due to differences in baseline fracture risk of patients identified by an FLS from all hip fractures or methods for fracture ascertainment.

Reduction of hip fracture risk depends on effective post fracture care to optimise functional recovery, reduce falls risk and improve bone health. This is further complicated by the limited evidence for falls prevention in patients with cognitive impairment[25]. In contrast, there is consistent evidence for the effectiveness of AOMs in patients with high fracture risk[26], post hip fracture[27] and with cognitive impairment[28]. The challenges for effective secondary fracture prevention using AOMs occur at the system, pharmacological, and patient level[9, 29]. At the system level, the patient pathway needs to reduce the time between fracture and initiation of therapy. This requires rapid identification by the FLS, brief assessments to determine fracture risk, and factors that may influence treatment choice, such as renal impairment, upper gastrointestinal disturbance, and previous AOM therapy. Bone mineral density is a recognised critical predictor of hip fracture risk[30] and is used to identify patients who would benefit from AOM. However, the place of DXA testing in patients who have already had a major osteoporotic fracture varies within and between countries. The UK’s National Institute for Health and Care Excellence (NICE) Technology Assessment 161 recommended a DXA scan may not be required if the responsible clinician considers it to be clinically inappropriate or unfeasible[3]. In 2019, the ASBMR recommended initiation of AOM in patients aged 65 years or over without the need for DXA if they had a hip or spine fracture[31]. During the recent COVID pandemic, there was a major impact on provision of DXA services worldwide[32] that led to recommendations to initiate AOM without DXA[33]. DXA is sometimes used to motivate patients to start and stay on treatment. Whilst a previous DXA scan has been shown to improve initial treatment adherence in younger individuals[34], the benefit in patients with a recent fragility fracture is unknown. DXA can be used to monitor treatment response; however, in the case of oral bisphosphonates, this has been shown to take more than 3 years[35] and so has limited value to improve adherence in the critical first years after an index fracture[10]. In patients who fracture on anti-resorptive therapy, there is a role for DXA to identify those patients who may be eligible for anabolic therapy in England and Wales as well as in many other countries. To reduce the time from fracture diagnosis to treatment initiation, a number of FLS models of care are now incorporating initiation of AOM within the trauma stay[36-38], particularly for patients requiring hospital admission because of their fracture. Whether the initiation of the AOM occurs within the acute trauma stay is not captured by the FLSDB.

Pharmacological aspects of the AOM include time to treatment onset and size of treatment fracture risk reduction. Bisphosphonates vary in their time to maximal fracture reduction with a weak effect from 4 months and a maximal effect requiring at least 12 months in general[39] with differences by fracture type and type of bisphosphonate[40]. A head-to-head trial with fracture end points has demonstrated the superiority of anabolic to anti-resorptive AOMs [41, 42]. In particular, anti-sclerostin therapy has been shown to rapidly increase hip bone density as measured by DXA or QCT faster and to a great extent than anti-resorptive or PTH analogues [43]. This is translated into substantial reductions in hip fracture risk within 12 months when treated with romosozumab compared to alendronate in women with the prior history of fracture, including with a recent hip fracture [42]. This is pertinent given the proportion of patients we observed with a subsequent hip fracture within a year of FLS identification. Finally, at the patient level, early initiation of treatment and support for adherence are required[44]. Adherence is poor with AOM therapy and improving adherence remains a substantial challenge for FLSs across the globe[45]. The use of parenteral therapies, requiring healthcare or homecare-based delivery, provide another potential approach to improve patient adherence[46].

The strengths of this study include its use of real-world data and hip fractures as an endpoint. A major limitation is that we were unable to adjust for confounding by indication. While we observed the proportion with a subsequent hip fracture was not lower than was seen in patients who an FLS decided did not need such medication, we recognise that patients recommended AOM therapy and different types of AOM therapy are likely to have different fracture risks, confounding by indication. For instance, those started on denosumab after their index hip fracture were older than those for whom an oral bisphosphonate was considered. However, choice between anti-resorptive therapies is usually guided by patient co-morbidities, likely survival and history of previous AOM use.. Further analyses using matching or adjustment methods to balance confounders are needed to examine whether those associations might be evidenced and to further our understanding of the benefits of AOM therapy in the FLS setting[47]. Secondly, there was a high level of missing data across the predictor variables, particularly for monitoring for treatment initiation and longer term adherence. This prevented additional scrutiny of AOM initiation and restricted our analysis to AOM recommendations. Another major limitation is that mortality data were not available so that the proportion of the missing follow up data explained by survival is unknown. In 2017, the average case identification rate for FLSs was 26.4% and further work is needed to examine if the increase in case finding since 2017 has altered the proportions having a subsequent hip fracture. This applies in particular to FLSDB records with an index hip fracture. In 2017, there were 64,642 hip fractures recorded in the NHFD[48], so the hip fractures recorded in the FLSDB in the same year represents a minority (15.3%) of all hip fractures with missing cases from non-participating (n=115/177) as well as participating hospitals. It is likely that the patients with hip fractures recorded in the FLSDB are not representative of hip fracture patients in general. as illustrated by the relatively high proportion referred for DXA as well as differences by age. Further work is needed to understand the patient and FLS service level characteristics of hip fractures that are submitted to the FLSDB including differences between FLSs. Due to the nature of the FLSDB, the specific site of fractures in the ‘other’ group are unknown, however, fractures of the face, skull, scaphoid, and digit are excluded in the guidance to users. More detailed differentiation of this group into major and minor fragility fractures may help identify additional patient groups with higher rates of subsequent hip fracture. Finally, it is important to note that in the absence of a control untreated arm, it is not possible to describe the impact of FLSs in reducing fracture risk vs. no FLS. However, a number of reviews have already demonstrated the effectiveness of FLSs compared with usual care[6, 7] and the aim of this analysis was to highlight the residual subsequent hip fractures after patients are seen by an FLS to inform FLS service improvement nationally.

In conclusion, using real-world data from a bi-national audit of FLSs, we have demonstrated a clinically important residual rate of subsequent hip fractures after FLS management. Rates of hip fractures were not lower by type of AOM recommended and further work is needed to better understand the reasons for this.

## Data Availability

All data produced in the present study are available upon reasonable request to HQIP

## Acknowledgements

MKJ was supported by the National Institute for Health Research (NIHR) Oxford Biomedical Research Centre (BRC). The views expressed are those of the author and not necessarily those of the NHS, the NIHR or the Department of Health.

## Supporting information

**S1 Fig.**
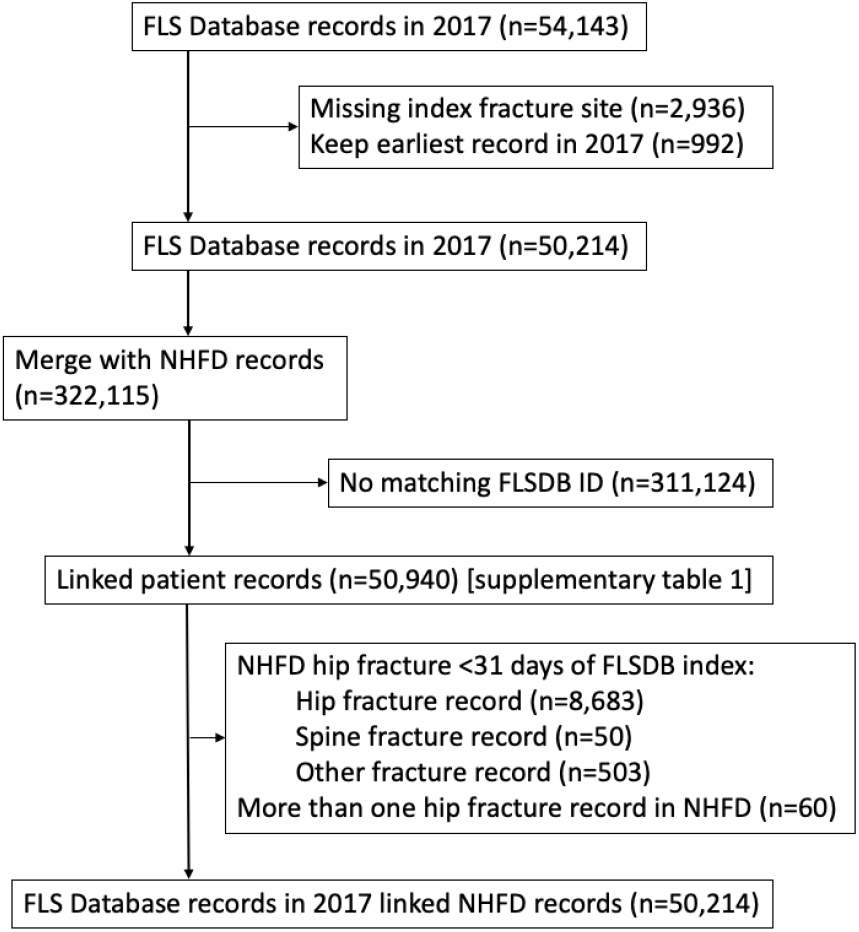
Linkage of FLSDB and NHFD cases.

**S1 Table:**
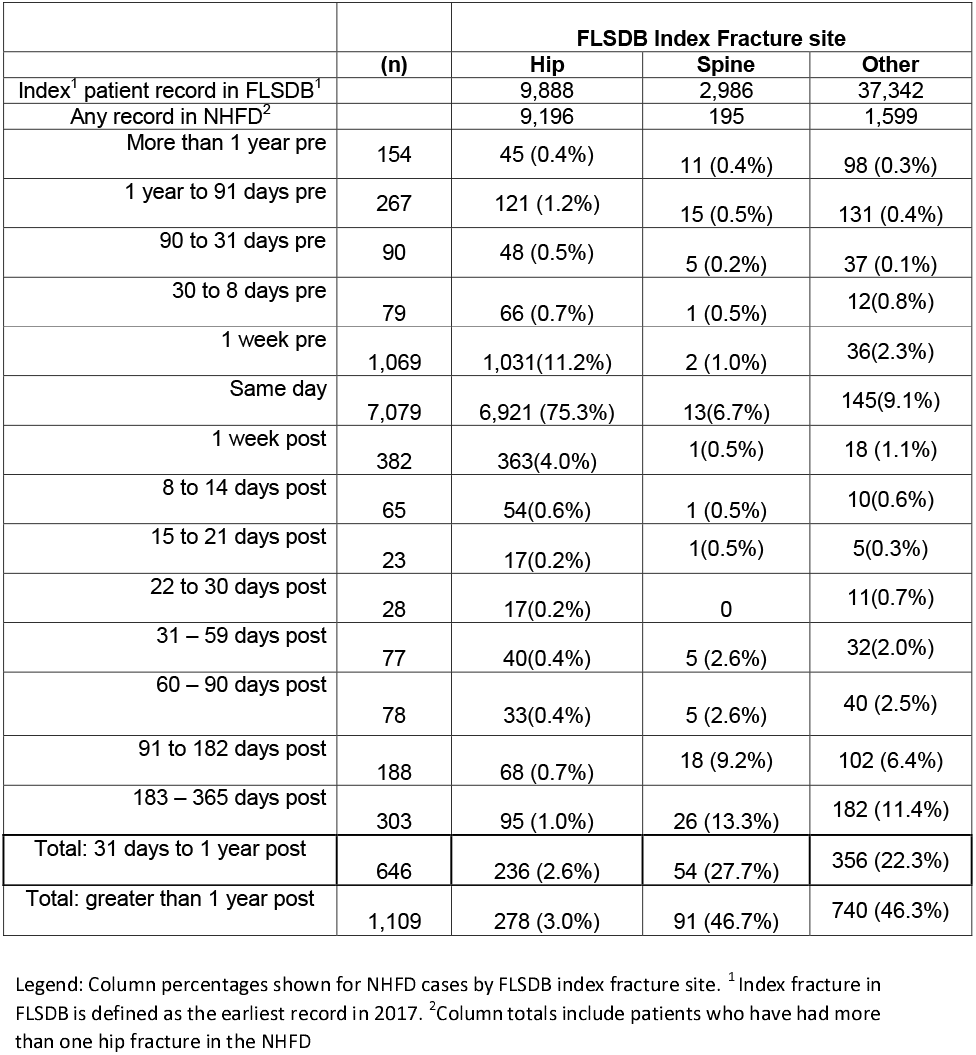
The difference in presentation date from the first fracture record from 1st January to 31st December 2017 in the FLSDB to a record in the NHFD from !st January 2016 to 31st December 2020 before removal of potential duplicates.

