## Supplementary Table 1 for "Subsequent hip fractures in patients identified by a Fracture Liaison Service (FLS) in England and Wales – linkage of the national FLS and Hip Fracture databases"

Supplementary material

**S1 Table: The difference in presentation date from the first fracture record from 1^st^ January to 31^st^ December 2017 in the FLSDB to a record in the NHFD from !st January 2016 to 31^st^ December 2020 before removal of potential duplicates.**

|  |  | **FLSDB Index Fracture site** | | |
| --- | --- | --- | --- | --- |
|  | **(n)** | **Hip** | **Spine** | **Other** |
| Index^1^ patient record in FLSDB^1^ |  | 9,888 | 2,986 | 37,342 |
| Any record in NHFD^2^ |  | 9,196 | 195 | 1,599 |
| More than 1 year pre | 154 | 45 (0.4%) | 11 (0.4%) | 98 (0.3%) |
| 1 year to 91 days pre | 267 | 121 (1.2%) | 15 (0.5%) | 131 (0.4%) |
| 90 to 31 days pre | 90 | 48 (0.5%) | 5 (0.2%) | 37 (0.1%) |
| 30 to 8 days pre | 79 | 66 (0.7%) | 1 (0.5%) | 12(0.8%) |
| 1 week pre | 1,069 | 1,031(11.2%) | 2 (1.0%) | 36(2.3%) |
| Same day | 7,079 | 6,921 (75.3%) | 13(6.7%) | 145(9.1%) |
| 1 week post | 382 | 363(4.0%) | 1(0.5%) | 18 (1.1%) |
| 8 to 14 days post | 65 | 54(0.6%) | 1 (0.5%) | 10(0.6%) |
| 15 to 21 days post | 23 | 17(0.2%) | 1(0.5%) | 5(0.3%) |
| 22 to 30 days post | 28 | 17(0.2%) | 0 | 11(0.7%) |
| 31 – 59 days post | 77 | 40(0.4%) | 5 (2.6%) | 32(2.0%) |
| 60 – 90 days post | 78 | 33(0.4%) | 5 (2.6%) | 40 (2.5%) |
| 91 to 182 days post | 188 | 68 (0.7%) | 18 (9.2%) | 102 (6.4%) |
| 183 – 365 days post | 303 | 95 (1.0%) | 26 (13.3%) | 182 (11.4%) |
| Total: 31 days to 1 year post | 646 | 236 (2.6%) | 54 (27.7%) | 356 (22.3%) |
| Total: greater than 1 year post | 1,109 | 278 (3.0%) | 91 (46.7%) | 740 (46.3%) |

Legend: Column percentages shown for NHFD cases by FLSDB index fracture site. ^1^ Index fracture in FLSDB defined as earliest record in 2017.

^2^Column totals include patients who have had more than one hip fracture in the NHFD
