## Supplementary figures and images for "Subsequent hip fractures in patients identified by a Fracture Liaison Service (FLS) in England and Wales – linkage of the national FLS and Hip Fracture databases"

### Supplementary figure 1

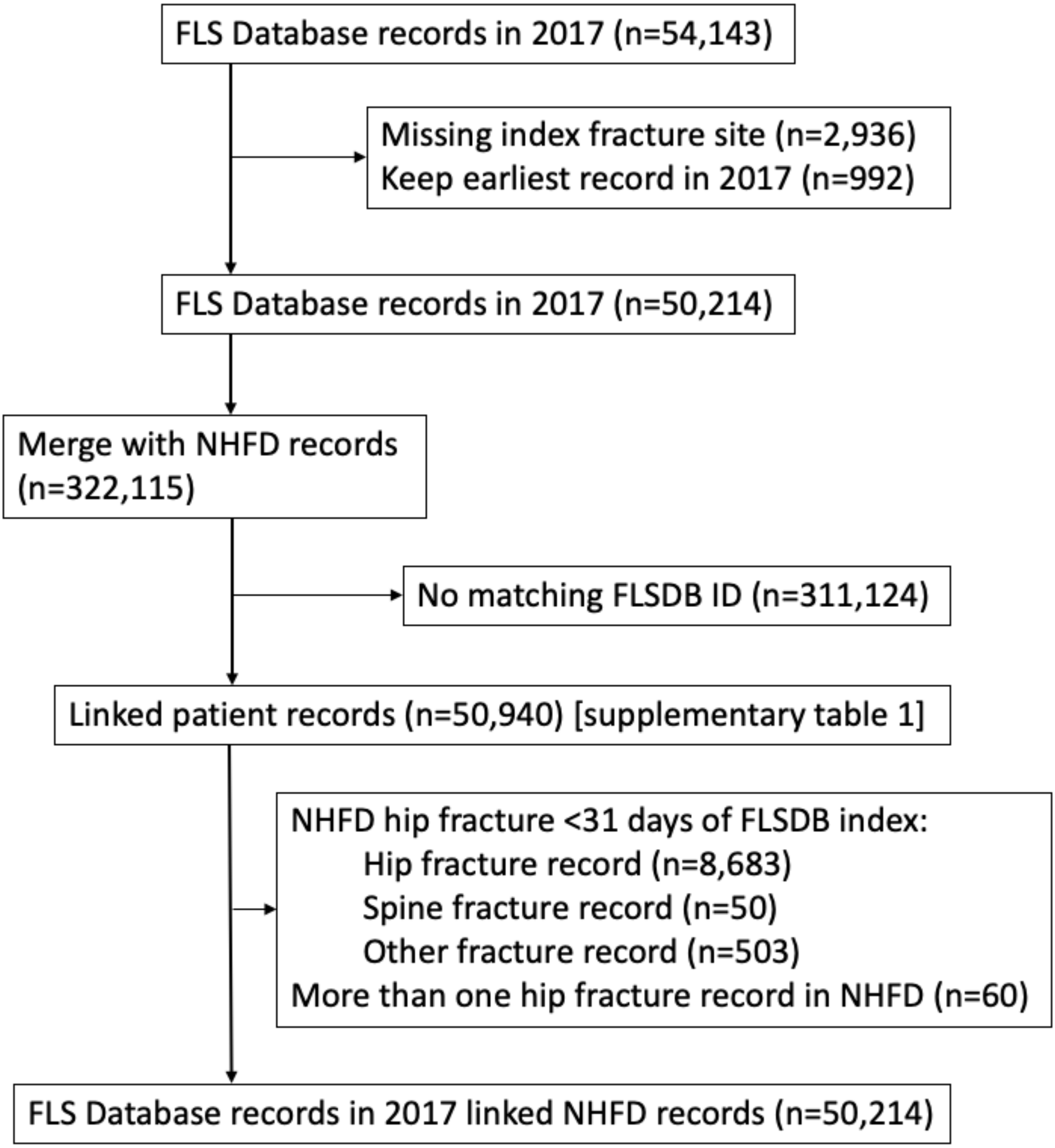
